# Genome-Wide Association Meta-Analysis Using a Recessive Model Illuminates Genetic Architecture of Type 2 Diabetes

**DOI:** 10.1101/2021.07.08.21258700

**Authors:** Mark J. O’Connor, Alicia Huerta-Chagoya, Paula Cortés-Sánchez, Silvía Bonàs-Guarch, Marta Guindo-Martínez, Joanne B. Cole, David Torrents, Kumar Veerapen, Niels Grarup, Mitja Kurki, Carsten F. Rundsten, Oluf Pedersen, Ivan Brandslund, Allan Linneberg, Torben Hansen, Aaron Leong, Jose C. Florez, Josep M. Mercader

**Author notes:** These authors jointly directed this work (Aaron Leong, Jose C. Florez, Josep M. Mercader). **Correspondence:** Josep M. Mercader, Ph.D., The Broad Institute of Harvard and MIT, 75 Ames Street, Cambridge, MA 02142.

## Abstract

**Objective:** Most genome-wide association studies (GWAS) of complex traits are performed using models with additive allelic effects. Hundreds of loci associated with type 2 diabetes have been identified using this approach. Additive models, however, can miss loci with recessive effects, thereby leaving potentially important genes undiscovered.

**Research Design and Methods:** We conducted the largest GWAS meta-analysis using a recessive model for type 2 diabetes. Our discovery sample included 33,139 cases and 279,507 controls from seven European-ancestry cohorts including the UK Biobank. We then used two additional cohorts, FinnGen and a Danish cohort, for replication. For the most significant recessive signal, we conducted a phenome-wide association study across hundreds of traits to make inferences about the pathophysiology underlying the increased risk seen in homozygous carriers.

**Results:** We identified 51 loci associated with type 2 diabetes, including five variants with recessive effects undetected by prior additive analyses. Two of the five had minor allele frequency less than 5% and were each associated with more than doubled risk. We replicated three of the variants, including one of the low-frequency variants, rs115018790, which had an odds ratio in homozygous carriers of 2.56 (95% CI 2.05-3.19, *P*=1×10^−16^) and a stronger effect in men than in women (interaction *P*=7×10^−7^). Colocalization analysis linked this signal to reduced expression of the nearby *PELO* gene, and the signal was associated with multiple diabetes-related traits, with homozygous carriers showing a 10% decrease in LDL and a 20% increase in triglycerides.

**Conclusions:** Our results demonstrate that recessive models, when compared to GWAS using the additive approach, can identify novel loci, including large-effect variants with pathophysiological consequences relevant to type 2 diabetes.

## INTRODUCTION

Type 2 diabetes affects nearly 1 in 12 adults globally (1), but its genetic architecture is still not fully understood. Over the last decade, large genome-wide association studies (GWAS) have used additive models to identify hundreds of associated loci (2–4). Additive models are most powerful when the effect of two copies of a risk allele is twice that of one copy. This model is computationally simple and statistically powerful, but it does not always match the pattern of inheritance of Mendelian disorders, including monogenic forms of diabetes, which can be transmitted in a dominant or recessive fashion (5). Variants with recessive effects, particularly low-frequency variants, can go undetected by additive models (6), suggesting that non-additive models have the potential to generate new biological insights.

To date, a handful of studies have used recessive models to identify genetic associations with type 2 diabetes, but these have been limited by small sample sizes (6–8). Nevertheless, some promising findings have emerged. In a Greenlandic population, homozygous carriers of two copies of a nonsense mutation in the *TBC1D4* gene, which facilitates glucose transfer into skeletal muscle in the setting of insulin stimulation, were found to have a tenfold increase in diabetes risk compared to other individuals in the same population (7). Hyperglycemia due to this variant occurs postprandially, so the diagnosis of type 2 diabetes in homozygous carriers often requires an oral glucose tolerance test, creating an opportunity for precision medicine (9). More recently, members of our group conducted GWAS with non-additive models for several age-related diseases (10) and identified multiple new loci, including one rare variant (rs77704739) associated with type 2 diabetes. This variant was also associated with reduced expression of the *PELO* gene, whose connection to diabetes is not well understood.

We have conducted the largest GWAS meta-analysis using a recessive model reported to date for type 2 diabetes. Over the last few years, GWAS sample sizes have grown exponentially (11), and reference panels for imputation have improved, making it easier to ascertain low-frequency variants accurately (12). To take advantage of these developments, we combined data from seven discovery cohorts and two replication cohorts to conduct the largest recessive-model GWAS yet reported for type 2 diabetes or any other disease. We identified and replicated multiple variants missed by larger additive studies, confirmed and fine-mapped the association near *PELO*, and conducted a phenome-wide association analysis to identify other impacted traits to better understand the pathophysiology underlying this novel association.

## RESEARCH DESIGN AND METHODS

### Study Population and Outcome Definition

We used data from multiple European-ancestry cohorts (Supplemental Table 1) including the UK Biobank (13), five cohorts known collectively as 70K for T2D (4), and the Mass General Brigham (MGB) Biobank (14). The UK Biobank is a sample of approximately half a million people recruited in the United Kingdom between the ages of 40 and 69 years. The 70K for T2D cohort consists of five studies with publicly available data, and the MGB Biobank consists of approximately 50,000 people recruited within a hospital system in the United States. We only considered individuals whose family relatedness was lower than that of third-degree relatives.

**Table 1:**
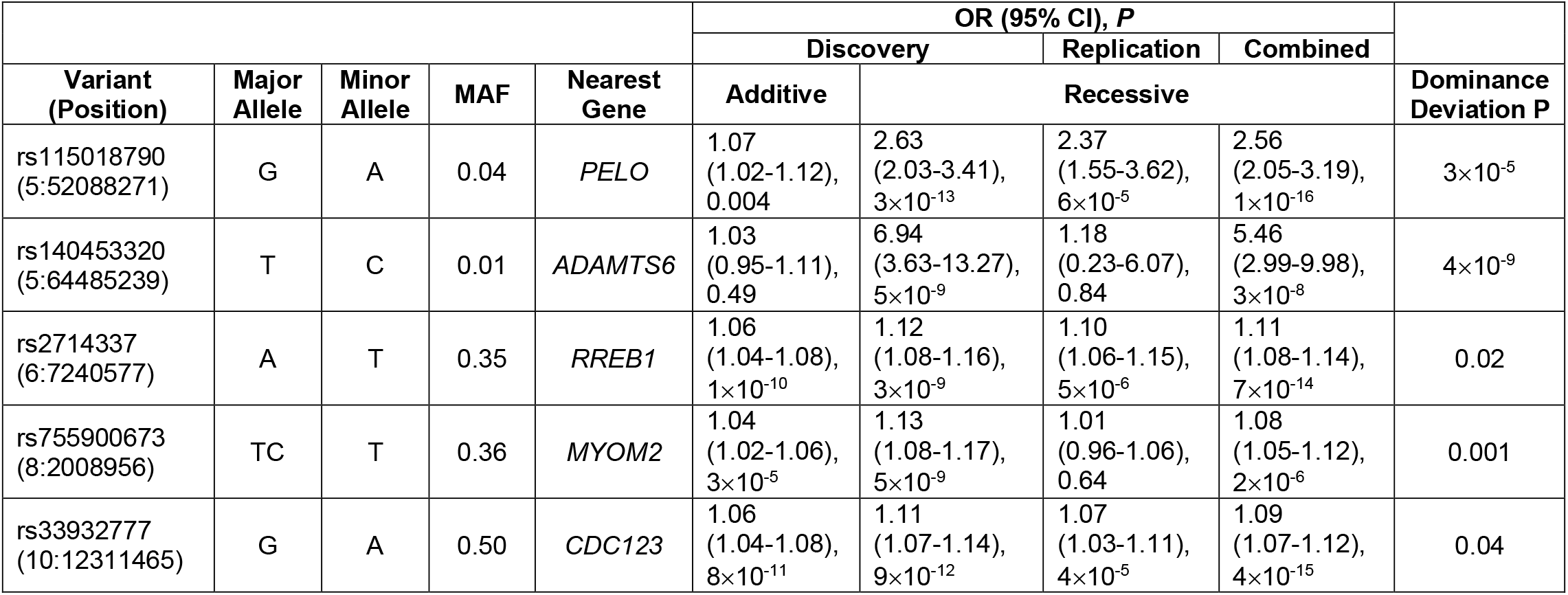
Novel recessively acting variants. Position is from genome assembly GRCh37 (hg19). For replication, variant rs140453320 was only assessed in FinnGen as there were only three homozygotes in the Danish cohort. Dominance deviation *P* values were calculated in the UK Biobank.

|  |  |  |  |  | OR (95% CI), *P* |  |  |  |  |
| --- | --- | --- | --- | --- | --- | --- | --- | --- | --- |
|  |  |  |  |  | Discovery |  | Replication | Combined |  |
| Variant<br>(Position) | Major Allele | Minor Allele | MAF | Nearest Gene | Additive | Recessive |  |  | Dominance Deviation P |
| rs115018790<br>(5:52088271) | G | A | 0.04 | *PELO* | 1.07<br>(1.02-1.12),<br>0.004 | 2.63<br>(2.03-3.41),<br>3×10<sup>-13</sup> | 2.37<br>(1.55-3.62),<br>6×10<sup>-5</sup> | 2.56<br>(2.05-3.19),<br>1×10<sup>-16</sup> | 3×10<sup>-5</sup> |
| rs140453320<br>(5:64485239) | T | C | 0.01 | *ADAMTS6* | 1.03<br>(0.95-1.11),<br>0.49 | 6.94<br>(3.63-13.27),<br>5×10<sup>-9</sup> | 1.18<br>(0.23-6.07),<br>0.84 | 5.46<br>(2.99-9.98),<br>3×10<sup>-8</sup> | 4×10<sup>-9</sup> |
| rs2714337<br>(6:7240577) | A | T | 0.35 | *RREB1* | 1.06<br>(1.04-1.08),<br>1×10<sup>-10</sup> | 1.12<br>(1.08-1.16),<br>3×10<sup>-9</sup> | 1.10<br>(1.06-1.15),<br>5×10<sup>-6</sup> | 1.11<br>(1.08-1.14),<br>7×10<sup>-14</sup> | 0.02 |
| rs755900673<br>(8:2008956) | TC | T | 0.36 | *MYOM2* | 1.04<br>(1.02-1.06),<br>3×10<sup>-5</sup> | 1.13<br>(1.08-1.17),<br>5×10<sup>-9</sup> | 1.01<br>(0.96-1.06),<br>0.64 | 1.08<br>(1.05-1.12),<br>2×10<sup>-6</sup> | 0.001 |
| rs33932777<br>(10:12311465) | G | A | 0.50 | *CDC123* | 1.06<br>(1.04-1.08),<br>8×10<sup>-11</sup> | 1.11<br>(1.07-1.14),<br>9×10<sup>-12</sup> | 1.07<br>(1.03-1.11),<br>4×10<sup>-5</sup> | 1.09<br>(1.07-1.12),<br>4×10<sup>-15</sup> | 0.04 |

Definitions of type 2 diabetes varied according to cohort. In the UK Biobank, for example, we used a validated algorithm designed specifically to identify cases of diabetes in that cohort (15). In the MGB Biobank, type 2 diabetes was defined according to an algorithm developed by the Biobank team (16) to have 99% positive predictive value. In the UK and MGB Biobanks, which both have a relatively low prevalence of type 2 diabetes, we excluded controls younger than 55 years, as the mean age of onset for type 2 diabetes is around 50 years (17).

### Recessive Genome-Wide Meta-Analysis

Genotyping, phasing, and imputation as well as sample and variant quality control were done according to cohort-specific protocols (Supplemental Table 1). For the recessive analysis in each cohort, we controlled for age, sex, body mass index (BMI), and principal components. For the UK Biobank, we also controlled for the genotyping platform, as two different genotyping arrays were used. For one of the five cohorts within 70K for T2D (6% of the cases in our discovery sample), age and BMI were not available. In our models, we used the minor allele in Europeans – not necessarily the non-reference allele – as the recessive allele to maximize our chances of identifying variants missed by prior GWAS.

For the UK and MGB Biobanks, computations were done using Hail version 0.2 (https://hail.is), and the 70K for T2D cohort was analyzed using the program SNPTEST (https://mathgen.stats.ox.ac.uk/genetics_software/snptest/snptest.html). After generating summary statistics using a recessive model for each cohort, we used the program METAL to meta-analyze the results (18), weighting cohorts by the inverse of the standard error for each variant. Our threshold for genome-wide significance was *P*=5×10^−8^, and we considered signals within 0.5 megabase pairs (Mb) to be part of the same locus. For comparison, we repeated our approach using an additive model. To visually inspect each genome-wide significant locus, we used the program LocusZoom (19). We estimated the power of our recessive and additive models to detect variants acting recessively across a range of allele frequencies and effect sizes using a simulation-based approach, assuming a baseline case prevalence of 10%, similar to our case-control ratio.

### Defining Novel Recessive Signals

We compared our results to the largest additive GWAS with available summary statistics (2, 3), and we defined signals as novel if they were not in significant linkage disequilibrium (LD) with a known signal (r^2^ < 0.3). This analysis was done using R version 3.6 (https://www.R-project.org) and the R package ‘LDlinkR’ (20, 21). The LD information was calculated using a British reference panel (1000 Genomes Project). For each signal, we used PLINK version 1.9 (22) to calculate a dominance deviation *P* value (23) using UK Biobank data. Signals were deemed to be non-additive if this *P* was less than 0.05. To ensure that signals near the major histocompatibility complex (MHC) region were not due to contamination of our cases with cases of type 1 diabetes, which is known to be heavily associated with haplotypes in the MHC region, we performed conditional analysis in the UK Biobank sample, adjusting for MHC haplotypes relevant to type 1 diabetes (24). We excluded variants that lost significance by more than one order of magnitude.

### Replication

We attempted to replicate our novel findings in two cohorts: FinnGen and a Danish cohort (Supplemental Table 2). FinnGen is a study based in Finland that combines genotyping with digital health data for over 100,000 people, and the Danish cohort consists of over 20,000 individuals (22% cases) from Denmark. The program SNPTEST was used to analyze both cohorts. We meta-analyzed the results from the replication cohorts with our initial results using the R package ‘rmeta’.

### Credible Sets

For each novel variant, we identified the set of variants with 99% probability of containing the causal variant. We used a Bayesian refinement approach (25), considering variants in LD with the lead variant (r^2^ > 0.1). Each credible set is akin to a confidence interval for the true causal variant. Within a locus, each variant is assigned an approximate Bayes factor (ABF) based on the following equation:

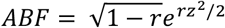

where *r* = 0.04/(*SE*^2^ + 0.04) and *z* = *β*/*SE*. The beta and standard error are the estimated effect size and corresponding standard error from the recessive-model logistic regression. This calculation assumes a Gaussian prior with mean 0 and variance 0.04. The posterior probability for a variant is equal to its ABF divided by the sum of all ABF values for the locus. Variants are ranked by ABF in decreasing order, and the cumulative probability is calculated starting at the top of the list and stopping when the value exceeds 99%.

### Colocalization with Gene Expression

To shed light on variants’ functional consequences, we used the Genotype-Tissue Expression (GTEx) project, version 8 (26). This database links genetic variants with tissue-specific gene expression, allowing for the identification of expression quantitative trait loci (eQTLs). For our most significant variant, which was an eQTL for the gene *PELO*, we performed colocalization analysis to confirm that our GWAS signal matched the signal influencing gene expression. Colocalization analysis compares *P* values for two traits across a locus to generate a posterior probability for the hypothesis that both traits are being influenced by the same variant. Due to the rarity of homozygous carriers of our variants, we used additive summary statistics from GTex for this analysis. We used the R package ‘coloc’ (27) and considered a window of 1 Mb around our leading signal.

### Phenome-Wide Association Study

For the variant located near the *PELO* gene, we performed a phenome-wide association study (PheWAS) in the UK Biobank, which provides detailed information about each participant’s health, dietary habits, and lifestyle characteristics. Phenotypes were curated and transformed using the PHEnome Scan ANalysis Tool, or PHESANT (28). As in our GWAS, we used a recessive model. We used logistic regression for binary phenotypes and linear regression for continuous phenotypes. We controlled for age, sex, ten principal components, and the genotyping platform. Limiting our binary phenotypes to those with more than five cases among homozygotes for the risk variant, we analyzed 1,731 binary phenotypes. We also analyzed 30 biomarkers such as cholesterol levels as well as 1,345 other continuous phenotypes. For significant associations, we used colocalization analysis to quantify the probability that the phenotype shared the same causal variant as type 2 diabetes. We also performed a PheWAS in the Danish cohort looking at 16 glycemic traits, using the same covariates as above.

### Sex-Stratified Analysis

To test whether the genetic effects of the variant near the *PELO* gene differed by sex, we performed a sex-stratified analysis within the UK Biobank for the biomarkers in our dataset and also for type 2 diabetes itself. We assessed the significance of the difference between sexes by including an interaction term in our regression model. We then confirmed sex-specific differences for type 2 diabetes in our two replication cohorts.

## RESULTS

### Genome-Wide Meta-Analysis Using a Recessive Model

Our discovery sample consisted of 33,139 cases of type 2 diabetes and 279,507 controls from seven cohorts. We meta-analyzed 11,634,328 variants and fitted additive and recessive models to compare the results. We identified 51 loci (Supplemental Table 3) that reached genome-wide significance in the recessive model, and 121 loci using the additive model (Figure 1). Of the 51 signals identified with the recessive model, 33% deviated from additivity (dominance deviation *P* < 0.05), and of these, five were distinct from the set of previously reported additive signals (Table 1).

**Figure 1:**
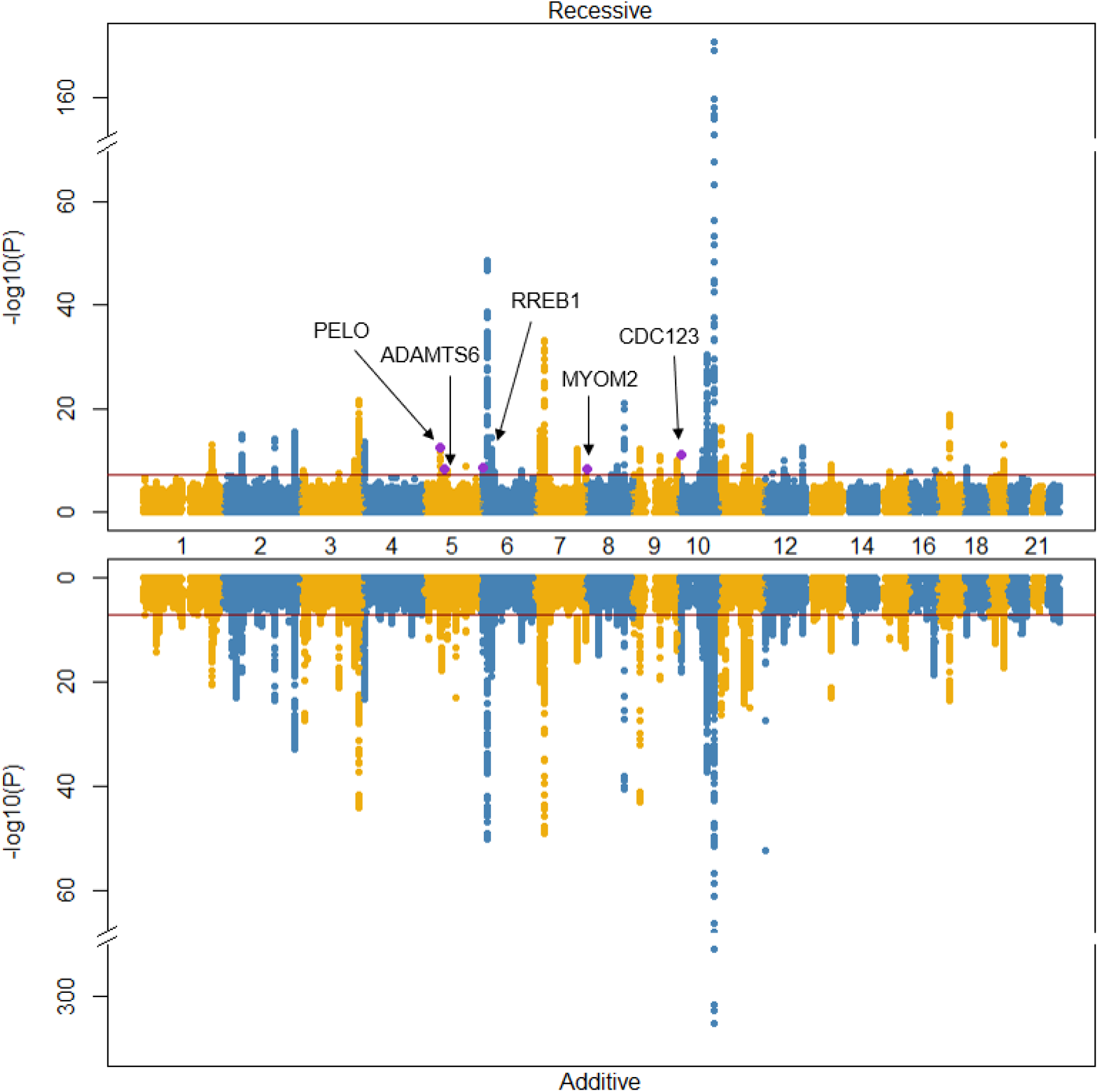
Miami plot comparing recessive and additive results. Non-additive signals are purple and labeled. The dark red line is the threshold for genome-wide significance.

The strongest recessive signal (rs115018790) was located within an intron of the *PELO* and *ITGA1* genes on chromosome 5 (Figure 2) and was in complete LD (r^2^ = 1) with the lead variant (rs77704739) that was previously identified in the GERA cohort (10), one of the discovery cohorts in this study. With minor allele frequency (MAF) 0.04, rs115018790 had an odds ratio (OR) for homozygous carriers of 2.63 (95% CI: 2.03-3.41), much greater than the additive-model OR of 1.07 (1.02-1.12). The *P* value for the recessive model (*P=*3×10^−13^) was ten orders of magnitude more significant than the additive one, and the dominance deviation test confirmed the variant’s recessive nature (*P*=3×10^−5^). This variant was near known additive signals (rs17261179, rs3811978, and rs62357230) associated with type 2 diabetes (2), but it was not in strong LD with any of these previously identified variants (maximum r^2^ = 0.08).

**Figure 2.**
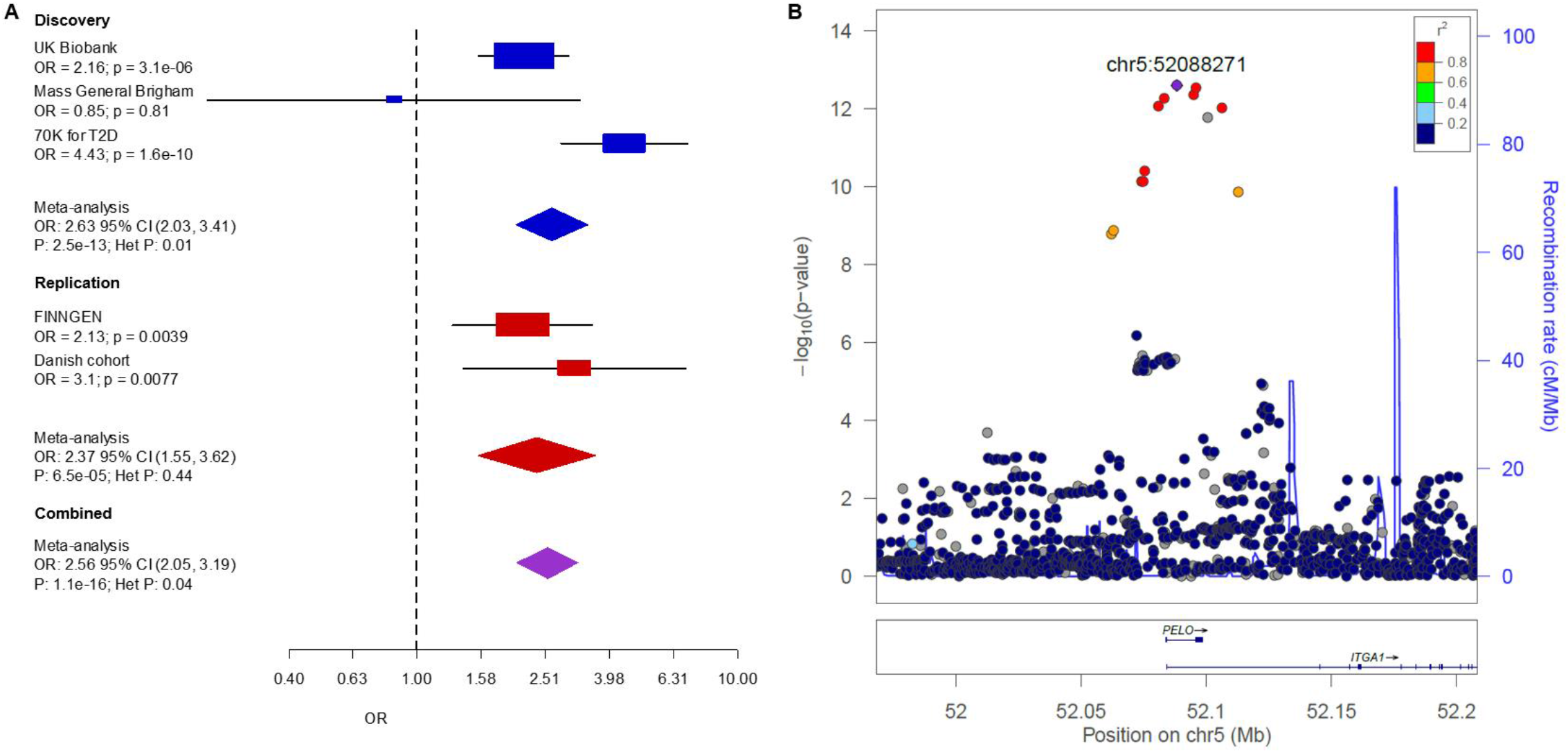
Replication of variant rs115018790. Panel A shows a forest plot of the discovery and replication cohorts. Cohort-specific odds ratios are denoted by boxes proportional to the size of the cohort, and error bars represent the 95% confidence interval. Panel B shows discovery GWAS *P* values at the *PELO* locus. Each dot represents a variant, with its genomic position (hg19) on the x axis and its *P* value (-log10) on the y-axis. Nearby genes are shown at the bottom of the plot.

We identified another non-additive, low-frequency variant (rs140453320) with large effect size on chromosome 5. This variant (MAF=0.01, OR [95% CI] = 6.94 [3.63-13.27], *P*=5×10^−9^) lies within an intron of the gene *ADAMTS6*. The additive *P* value was 0.48, leading to highly significant dominance deviation (*P*=4×10^−9^). This signal was over 1 Mb away from any previously known signal associated with type 2 diabetes.

The other three novel non-additive signals were significantly more common, each with MAF > 30%. Two of the three were located less than 0.5 Mb from known additive loci, but these signals were in weak LD with previously reported associations, with maximum r^2^ between 0.1 and 0.3 (Supplemental Table 4). The third (rs755900673) was an indel (OR [95% CI] = 1.13 [1.08-1.17], *P*=5×10^−9^) on chromosome 8 located within an intron of the *MYOM2* gene, more than 7 Mb away from any locus additively associated with type 2 diabetes.

We performed power simulations for our top variant (rs115018790, MAF 0.04, OR 2.63) and found that a GWAS with an additive model with our case-control ratio would need approximately 1.8 million participants to have 80% power to detect a genome-wide significant signal whereas a recessive model would only need 160,000 participants. At higher allele frequencies, the benefits of the recessive model become much less pronounced (Supplemental Figure 1).

### Replication

Our two replication cohorts consisted of 28,336 cases and 62,253 controls. Of the four non-additive signals for which we had sufficient power, three replicated, and one did not (Table 1, Supplemental Figure 2). Variant rs115018790 replicated in both cohorts (meta-analysis OR [95% CI] = 2.56 [2.05-3.19], *P*=1×10^−16^).

Two of the other three variants for which we had power also replicated. The indel near *MYOM2*, rs755900673, did not replicate (*P*=0.84) and showed high heterogeneity (*P*=0.008). Our power to replicate the rare variant near *ADAMTS6* was limited because there were only 10 homozygous carriers in our replication sample compared to 74 in our discovery sample. This signal retained genome-wide significance when we meta-analyzed the discovery and replication cohorts (*P*=3×10^−8^).

### Gene Expression Colocalization Analysis

Using GTEx data (26), we found that rs115018790 was associated with reduced *PELO* expression in multiple tissues. Colocalization analysis, which tests the hypothesis that traits are associated and share a single causative variant, confirmed the link between rs115018790 and reduced *PELO* expression in many tissues, including subcutaneous adipose tissue (posterior probability 0.99, *n*=581), skeletal muscle (0.99, *n*=706), and the pancreas (0.96, *n*=305). Colocalization plots (Supplemental Figure 3) comparing the recessive *P* values for the association with type 2 diabetes to additive *P* values from the gene expression dataset showed a high degree of correlation between the two sets of *P* values, visually confirming rs115018790’s connection to reduced *PELO* gene expression. The signal’s 99% credible set (Supplemental Table 5) contains rs185240714 (posterior probability 0.23), which is located in the 5’ untranslated region of the *PELO* transcription start site, further supporting a causal link.

### Phenome-wide Association Study

Using a recessive model, we found that multiple biomarkers (Figure 3) were associated with rs115018790 in the UK Biobank. Homozygotes for the risk allele had significantly higher triglycerides and lower LDL, HDL, and total cholesterol. Effect sizes were large. For example, being a homozygous carrier of the risk allele was associated with a 0.35 mmol/L (14 mg/dL) decrease in LDL (10% change relative to the mean) and a 0.35 mmol/L (31 mg/dL) increase in triglycerides (20%). These associations, particularly for triglycerides, were less significant using an additive model (Supplemental Table 6), suggesting that rs115018790 acts in a recessive manner for these traits as well. Colocalization analysis (Supplemental Figure 3) confirmed that these lipid associations are the result of a single shared variant (posterior probability > 0.99 for each trait). It did not appear that medication use was responsible for the observed effects on lipids because homozygotes for the risk allele were less likely (OR [95% CI] = 0.66 [0.49-0.88], *P*=0.005) to be on LDL-lowering therapy. Other biomarkers associated with rs115018790 included albumin and C-reactive protein. There was also a nominally significant (*P*=0.01) association with estradiol. The other novel variants did not have comparably significant and numerous biomarker associations (Supplemental Table 7).

**Figure 3:**
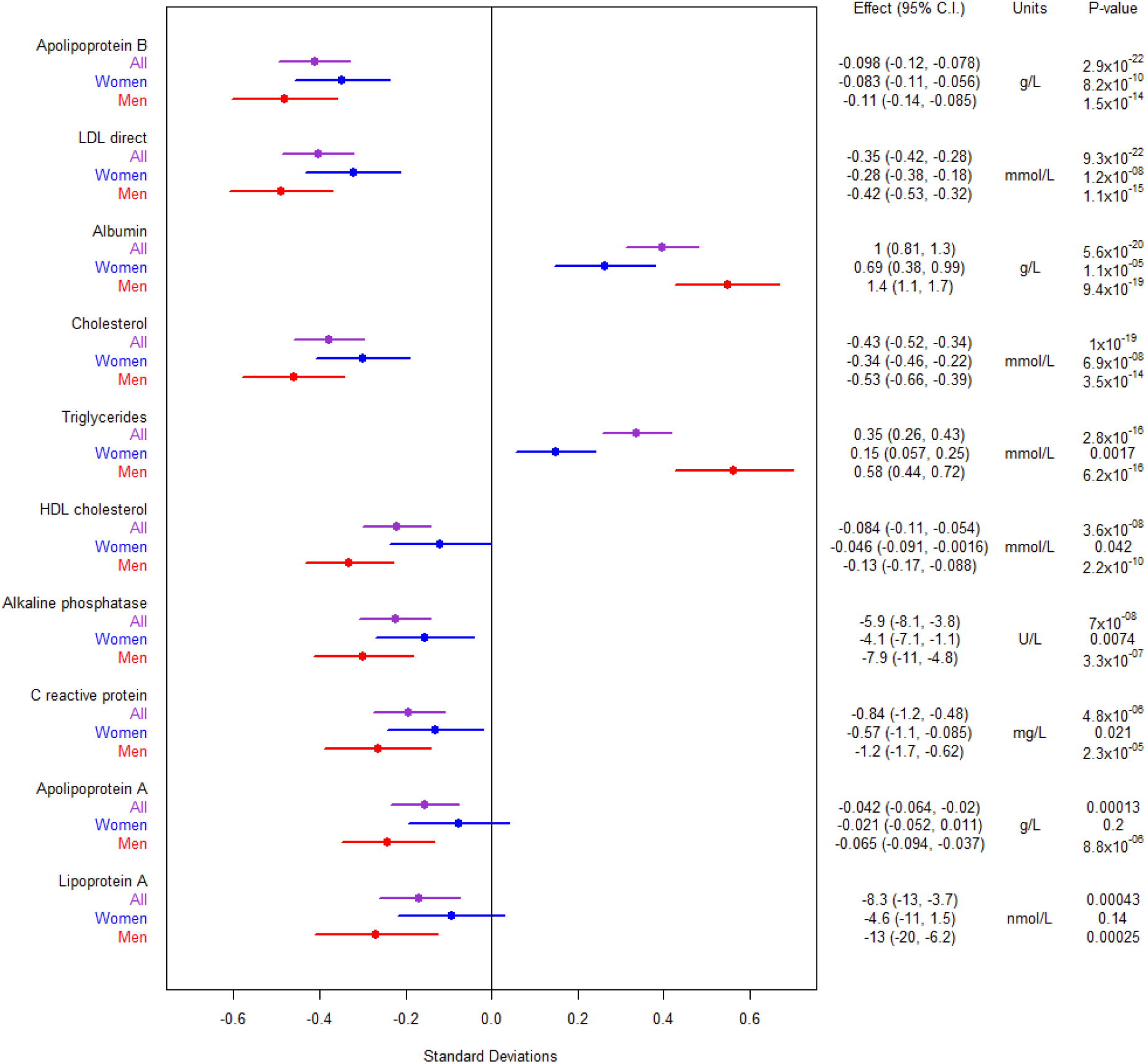
Biomarker associations for variant rs115018790. The figure to the left shows effect sizes normalized by each trait’s standard deviation. The error bars in the figure to the left represent 95% confidence intervals.

When we examined non-biomarker phenotypes (Supplemental Table 8) using a recessive model, we found that the variant near *PELO* was associated with a variety of hematologic features (decreased blood cell count, increased reticulocyte count and percent, increased mean corpuscular hemoglobin and volume, and decreased red blood cell distribution width) as well as increased alcohol intake frequency. None of the binary phenotypes reached a strict Bonferroni-corrected significance threshold of 1.6×10^−5^, but the top two phenotypes were metformin use (OR [95% CI] = 2.27 [1.56-3.31], *P*=2×10^−5^) and “diabetes diagnosed by a doctor” (1.87 [1.39-2.54], *P*=6×10^−5^). We did not detect significant associations with cardiovascular phenotypes such as heart attack or stroke after correcting for multiple testing.

In the Danish cohort, our power to detect recessive associations with glycemic traits was limited due to the low number of homozygous carriers (Supplemental Table 9). None of the traits were recessively associated with the variant. In an additive analysis, the variant was associated with increased insulin and C-peptide levels at the 120-minute timepoint of an oral glucose tolerance test.

### Sex-stratified Analysis

Because the variant near *PELO* was nominally associated with estradiol, we performed an analysis stratified by sex and, in the case of women, by menopause status, and we found that the effect of rs115018790 on estradiol was only significant in pre-menopausal women, with homozygotes for the risk allele having higher estradiol levels (174 pmol/L [95% CI: 49-300], *P*=0.006) than other pre-menopausal women. For other biomarkers such as cholesterol and triglycerides, effects were stronger in men than in women (Figure 3, Supplemental Table 6). For example, the recessive association with triglycerides was over twelve orders of magnitude more significant in men (*P*=3×10^−16^) than in women (*P*=0.002).

We also performed a sex-stratified analysis for type 2 diabetes itself. In the UK Biobank, we found that the association was limited to men (OR [95% CI] = 3.05 [2.10-4.43], *P*=5×10^−9^) as opposed to women (0.89 [0.42-1.87], *P*=0.75), with an interaction *P* value of 7×10^−7^. We replicated this finding in our replication cohorts (Supplemental Table 10). Meta-analysis across cohorts confirmed a large effect in men (OR=2.99 [2.18-4.10], *P*=1×10^−11^) and little to no effect in women (OR=1.41 [0.87-2.28], *P*=0.15).

### Comparison with Known Lipid-Associated Variants

To put effect sizes into context, we compared rs115018790 to previously described lipid-related variants of large effect size (29) using UK Biobank data. The LDL-lowering effect (0.35 mmol/L or 14 mg/dl) of rs115018790 in homozygotes was comparable to the effect (0.34 mmol/L or 13 mg/dl) of carrying one copy of a well-known protective variant (rs11591147) associated with the *PCSK9* gene. The triglycerides-increasing effect (0.35 mmol/L or 31 mg/dl) of rs115018790 in homozygotes was larger than the change (−0.29 mmol/L or −26 mg/dl) seen in homozygotes for a known variant (rs1569209) linked to the lipoprotein lipase (*LPL*) gene, known to be involved in triglyceride metabolism. For men, the size of rs115018790’s effect (0.58 mmol/L or 51 mg/dl) was almost double that of the *LPL* variant.

## DISCUSSION

Type 2 diabetes is a highly polygenic trait, and hundreds of loci associated with the disease have been identified, mostly via large GWAS meta-analyses conducted under additive genetic models (2, 3). This prior work has produced useful results, identifying potential therapeutic targets and also allowing for the creation of polygenic scores capable of quantifying one’s genetic risk (30). A sizeable fraction of the heritability of type 2 diabetes, however, remains unexplained by loci identified using additive models. Recessive modeling offers a way to identify new associations, creating opportunities for discovery and improved genetic risk stratification.

Our work takes advantage of the increasing number of genetic datasets now available, and it is currently the largest GWAS using a recessive model yet reported for type 2 diabetes or any other complex disease. We were able to identify multiple variants acting recessively, including two low-frequency variants of large effect size. Most of the variants identified via additive analyses have ORs less than 1.1, but the most significant variant we identified had an OR of 2.56 in homozygous carriers. Our minimum sample size to detect this variant was ten times smaller because we used a recessive, not an additive, model.

This variant was located near the *PELO* gene, and one of the six variants in the 99% credible set was in the gene’s upstream 5’ untranslated region, suggesting a role for this variant in gene expression regulation, a link we confirmed across multiple tissues using colocalization approaches. Members of our group first identified this association in one of our cohorts while conducting recessive-model GWAS for multiple age-related diseases (10). In this study, we confirmed the association with a larger sample size, fine-mapped the region, and used the power of the UK Biobank to demonstrate that the phenotypic effects of this variant are not limited to type 2 diabetes.

Homozygous carriers of the *PELO* variant exhibit significantly different circulating triglyceride and cholesterol levels compared to other individuals. These effects were most pronounced in men but were also seen in women, and the effect sizes were clinically relevant and comparable to previously discovered genetic variants that revealed novel therapeutic targets. The reduction in LDL associated with rs115018790 was approximately 10% (given an average LDL of 3.62 mmol/L or 140 mg/dl) whereas statins, the most commonly used LDL-lowering medications, typically lower LDL by 30 to 60% (31). As would be expected for carriers of an LDL-lowering variant, homozygotes for the minor allele at rs115018790 were less likely to be on statin medication. For triglycerides, the effect size (20%) was even larger.

The overall consequences of the effect of variant rs115018790 on lipid levels remain unclear. Low LDL is known to protect against cardiovascular events. High triglycerides and low HDL, on the other hand, are associated with cardiovascular disease, although for these two lipid particles, it is not clear whether the relationship is causal (32, 33). For homozygotes at rs115018790, the protective effects of lower LDL may be offset by the high triglycerides and lower HDL, meaning that the net effect on cardiovascular risk could be beneficial, harmful, or neutral. Our PheWAS did not reveal associations with cardiovascular events such as myocardial infarction or stroke. This lack of association, however, must be interpreted with caution in the setting of limited power, automatically curated phenotypes, and “healthy volunteer” selection bias in the UK Biobank (34).

The mechanism by which *PELO* affects diabetes risk is not clear. The gene is ubiquitously expressed, and its genetic deletion in mice leads to embryonic lethality (35). It is evolutionarily conserved and plays a role in rescuing stalled ribosomes, thus affecting the translation of multiple mRNA transcripts (36). It has a known role in sustaining protein synthesis in developing blood cells and platelets (37). A recent CRISPR loss-of-function screen in human pancreatic beta cells suggests that *PELO* may also play a role in insulin secretion (38). The sex-specific effect on diabetes risk in our study was striking, and more investigation is needed to determine what factors underlie the increased risk in men.

One limitation of our study is the restriction of the analyses to participants of European ancestry. Estimation of recessive effects requires large sample sizes, as homozygous carriers of low-frequency variants are rare. Progress has been made in terms of recruiting diverse participants for genetic studies, but people of European ancestry still make up the bulk of available datasets. In the future, non-additive methods may yield new insights when applied to non-European populations, work that could be particularly fruitful given the increased genetic diversity of these populations (39) and the increasing availability of multi-ethnic cohorts (40).

It is worth noting that most of the associations detected in our recessive analysis had already been uncovered in prior additive GWAS. This observation matches our power simulations comparing additive and recessive models. In these simulations, the benefit of the recessive model was significant at the low end of the allele-frequency spectrum whereas both models had similar power to detect high-frequency variants with recessive effects.

Our work illustrates the value of performing non-additive analyses to uncover low-frequency recessive variants. By conducting what is currently the largest GWAS using a recessive model for type 2 diabetes, we confirmed that a variant linked to reduced *PELO* gene expression appears to have significant effects not just on diabetes but also on lipid metabolism. Recessive models of type 2 diabetes and glycemic traits as part of larger and more diverse genetic discovery efforts are likely to provide additional associations that will in turn provide a better understanding of diabetes pathophysiology.

## Data Availability

The data supporting the findings of this study are available within the article and its supplementary materials. Additional data are available from the corresponding author J.M.M. upon reasonable request.

## Acknowledgements

We acknowledge Bianca C. Porneala, M.S., for technical assistance in the collection and curation of the genotype and phenotype data from the MGB Biobank. We also acknowledge Josee DuPuis, Ph.D., and Peitao Wu, Ph.D., for their valuable feedback.

## Author Contributions

J.M.M. and A.L. planned the analysis. M.J.O. performed the GWAS in the UK Biobank and MGB Biobank. A.H.-C. and J.B.C assisted M.J.O. with these cohorts. P.C.S., S. B.-G., D.T., and J.M.M. analyzed the 70K for T2D cohort. K.V. performed and M.K. supervised replication analysis in FinnGen, and C.F.R. performed the analysis in the Danish cohort supervised by N.G. and T.H. and with assistance from O.P., I.B., and A.L., who all contributed to assembling the data. M.J.O. performed the meta-analysis and the PheWAS. M.J.O. and J.M.M. wrote the manuscript. J.M.M., A.L., and J.C.F. supervised the work. J.M.M. is the guarantor of this work and, as such, had full access to all of the data in the study and takes responsibility for the integrity of the data and the accuracy of the data analysis.

## Prior Presentation

Parts of this work were presented in an abstract as part of the American Diabetes Association’s 80^th^ Scientific Sessions, June 2020.

