## Supplemental Figure for "Genome-Wide Association Meta-Analysis Using a Recessive Model Illuminates Genetic Architecture of Type 2 Diabetes"

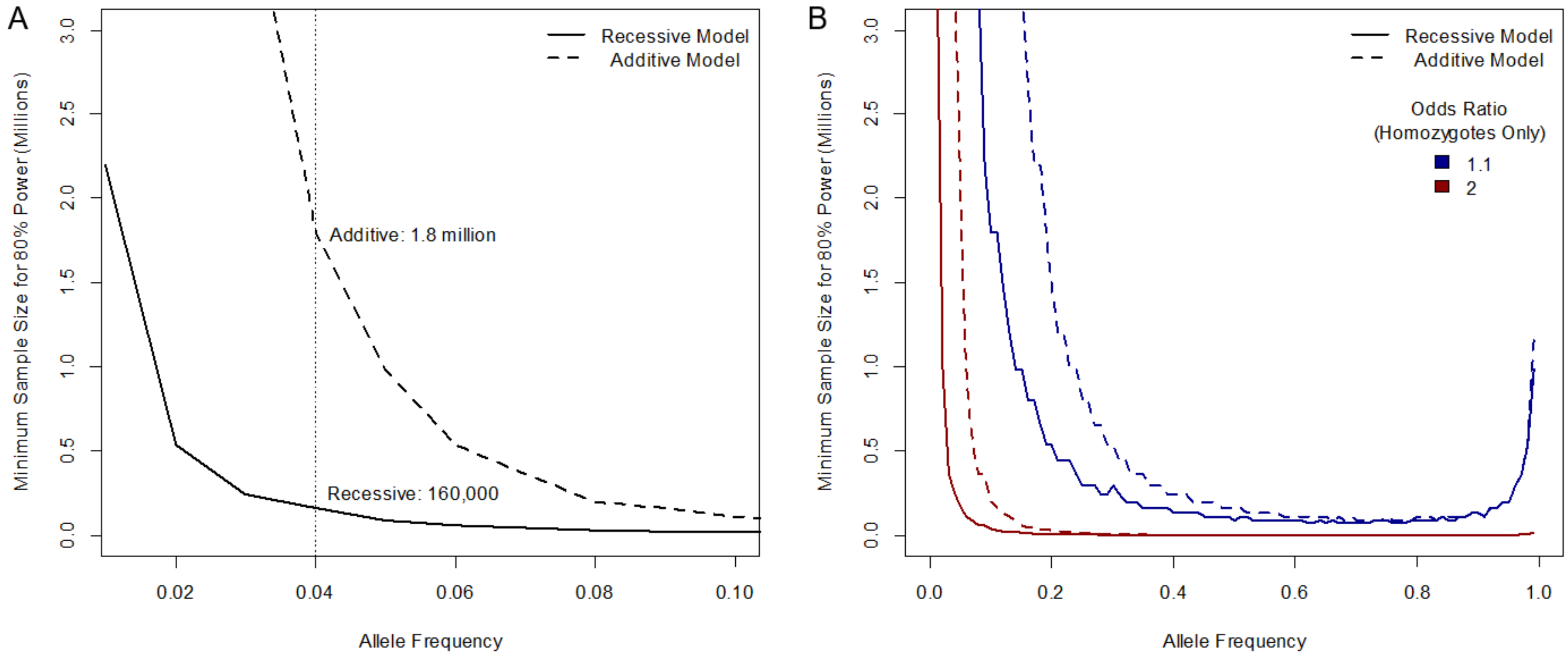


**Supplemental Figure 1: Power simulation comparing additive and recessive model.** Panel A shows results for a recessively acting variant with OR 2.63, equal to that of variant rs115018790 in the discovery cohort. The vertical dotted line represents the actual minor allele frequency (0.04) of rs115018790. Panel B shows results at the low end of the allele-frequency spectrum for two different ORs. Lines for recessive models are solid, and for additive, dashed.

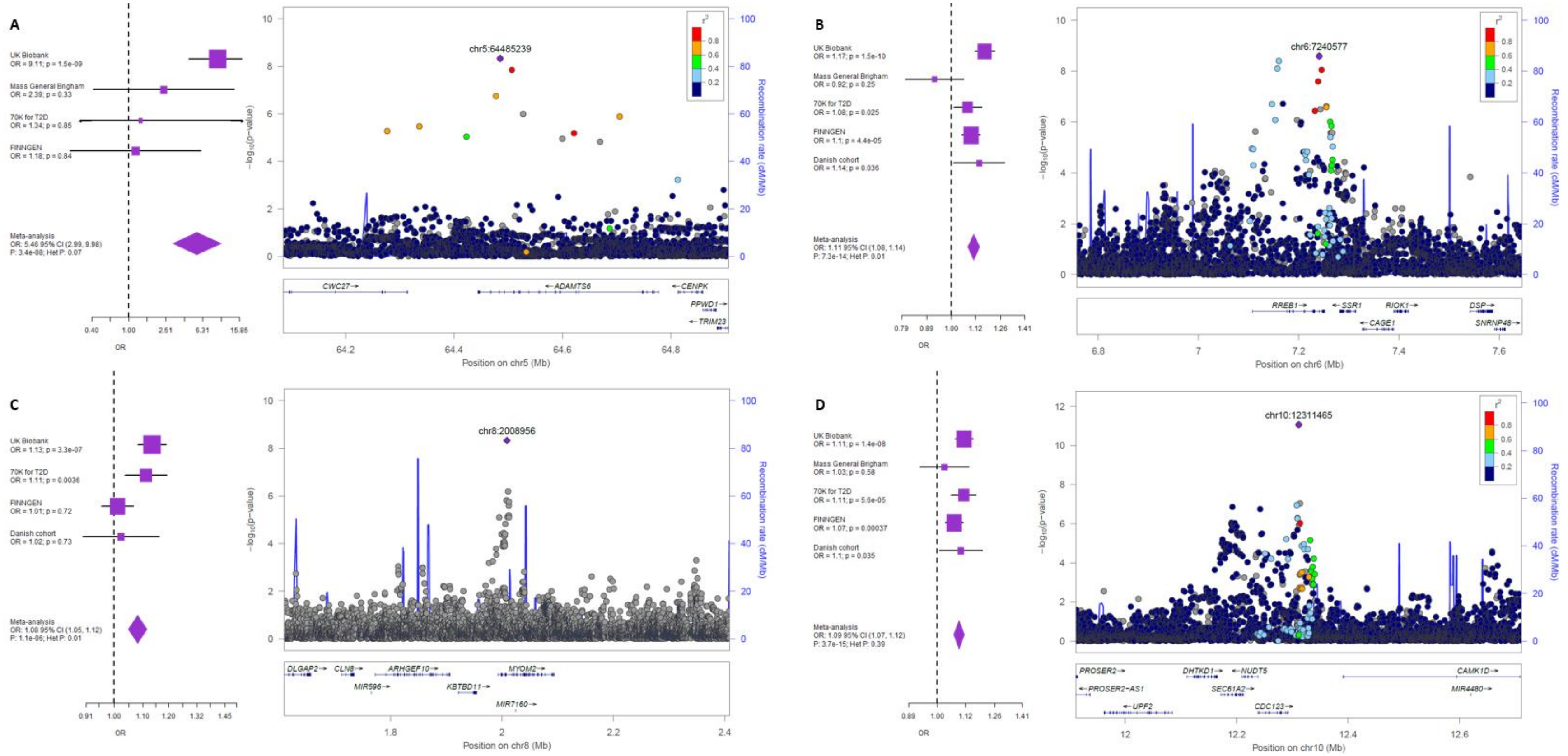


**Supplemental Figure 2: Additional non-additive variants.** Each panel shows a forest plot and colocalization plot for one the following variants: rs140453320 (panel A), rs2714337 (B), rs755900673 (C), and rs33932777 (D). In each forest plot, cohort-specific odds ratios are denoted by boxes proportional to the size of the cohort, and error bars represent the 95% confidence interval. Each colocalization plot shows discovery GWAS *P* values, with each dot representing a variant with the genomic position (hg19) on the x axis and the *P* value (-log10) on the y-axis. LD information was not available for rs755900673, an indel.

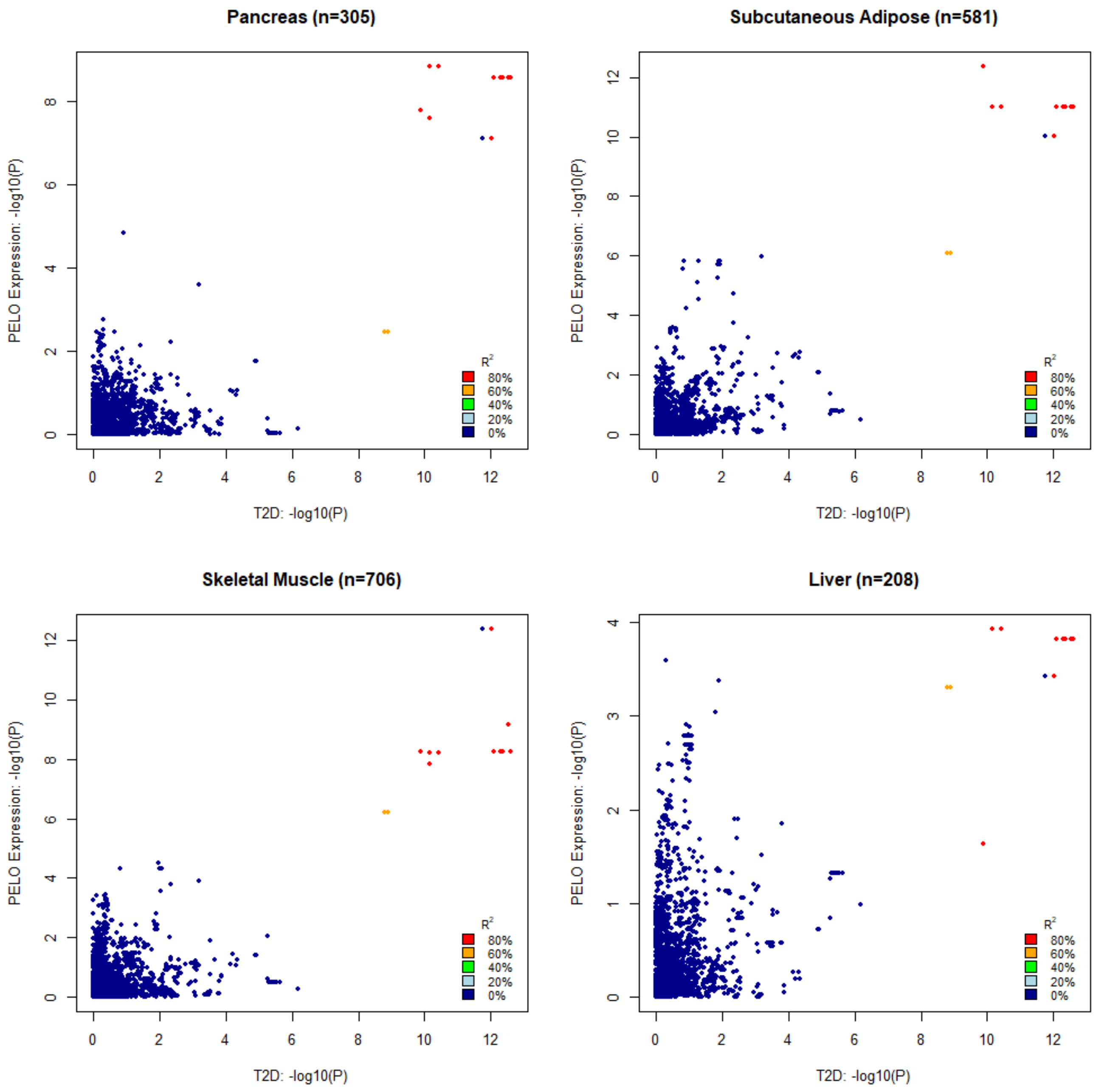


**Supplemental Figure 3: Colocalization plots for variant rs115018790 and *PELO* expression across tissues.** The *PELO* expression *P* values were calculated using an additive model due to the limited tissue-specific sample sizes.

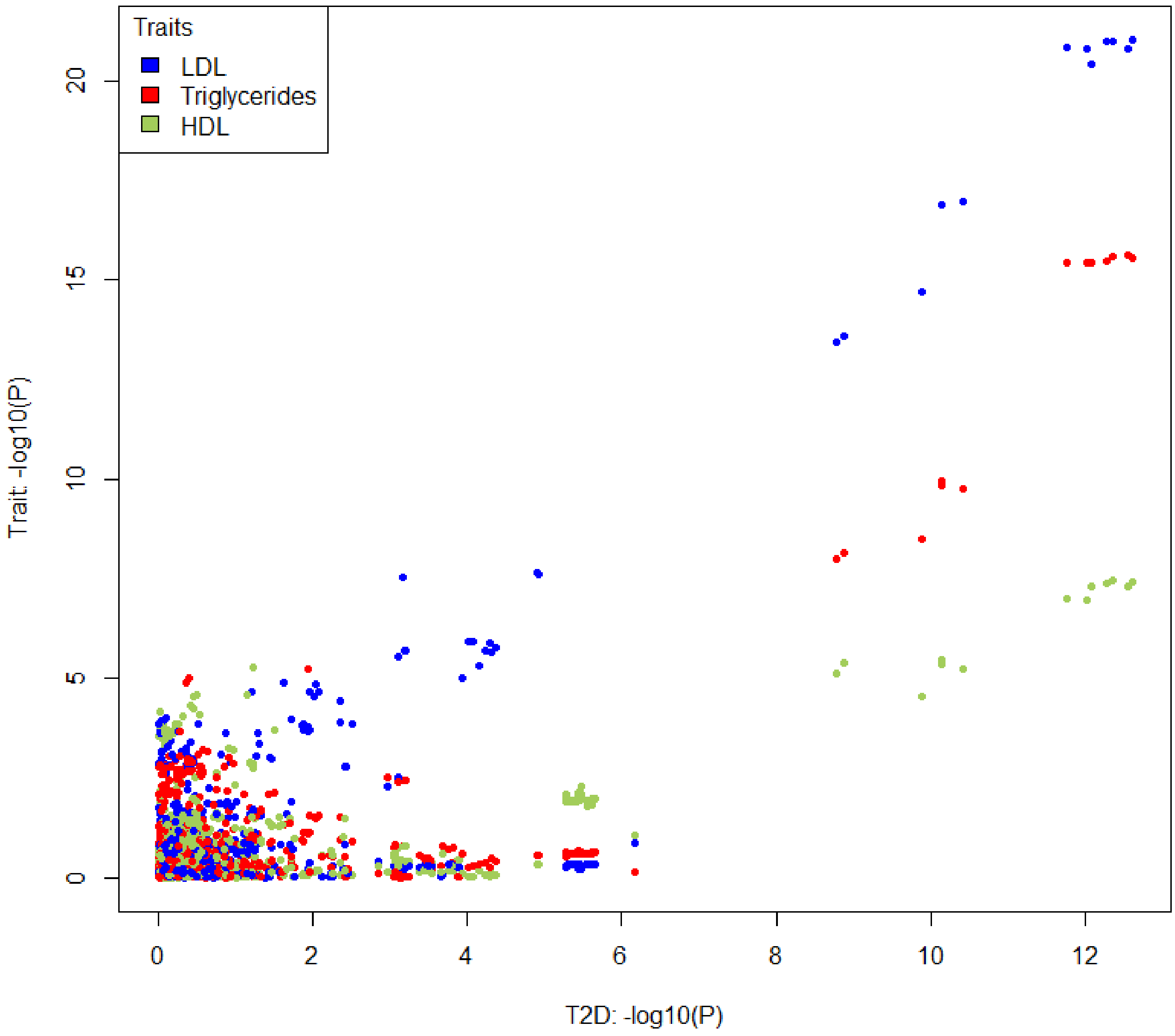


**Supplemental Figure 4: Colocalization plot for variant rs115018790 and lipid levels.** All models were recessive. The posterior probability of one causal variant for both traits was over 99% for each of the three lipid levels.
